# VectBox Sync: a web application for vectorcardiographic analysis of ventricular synchrony from 12-lead electrocardiograms

**DOI:** 10.64898/2026.11.09.26367663

**Authors:** Ramiro Goldman, Damian Spagnuolo, Osvaldo Chara, Juan Fernández

## Abstract

We introduce VectBox Sync, a browser-based application that implements a published vectorcardiographic (VCG) dyssynchrony index for exploratory assessment of ventricular electrical synchrony from the 12-lead electrocardiogram (ECG). Despite guideline-directed patient selection, a substantial fraction of patients who receive cardiac resynchronization therapy (CRT) do not obtain the expected benefit. QRS duration and morphology remain the principal electrical criteria, yet they do not capture the three-dimensional trajectory of ventricular activation. VectBox Sync prioritizes native digital ECG recordings, including matrices and common clinical and research waveform formats; optional manufacturer-PDF import and externally digitized numerical files extend access when paper-like archives are the only available source. After optional preprocessing, the user selects a QRS interval, and the application calculates the dyssynchrony index relative to a calibrated control cutoff. It also provides companion VCG descriptors (voltage and speed integrals, loop angles, and planarity), together with VCG trajectories, synchrony-field maps, and a multilingual PDF report. Labeled conditions from the same session can be overlaid for comparison, for example across successive pacing settings. VectBox Sync makes this VCG-based assessment accessible through an integrated web application, providing quantitative descriptors that complement conventional ECG criteria. The application is available at https://vectbox.com/sync for research and educational use.

## 1 Introduction

Ventricular electrical dyssynchrony is a key factor in cardiac remodeling and in predicting response to cardiac resynchronization therapy (CRT). Despite guideline-directed selection, a sub-stantial proportion of patients undergoing CRT do not achieve the expected clinical benefit[1]. Beyond QRS duration and morphology on the standard 12-lead electrocardiogram (ECG), vectorcardiographic (VCG) descriptors summarize the spatial evolution of ventricular activation and can support quantitative synchrony assessment[2], including the three-dimensional QRS area[3, 4] and other metrics linked to CRT response[5]. Under normal conduction, ventricular depolarization is a relatively conserved space-time process: differences in the amount of electrically involved myocardium mainly change the scale of the cardiac vector rather than its activation trajectory. Voltage-time and speed-time integrals of that vector (VTI and STI) are then expected to co-scale, so their relationship is approximately preserved. Conduction disease changes the sequence of activation, not merely its amplitude; VTI and STI can then change differentially, and we proposed the deviation from the normal VTI–STI relationship as a dyssynchrony index: a global descriptor of that organizational change, complementary to QRS duration[2].

That index was computed in our previous work with an internal analysis pipeline. **VectBox Sync** is a hosted web technology that makes the same workflow usable outside our laboratory, without local software installation. Its present use is exploratory research and teaching in that clinical setting: load a digital 12-lead ECG, select a QRS segment interactively, obtain the index and companion VCG descriptors with visualization, optionally compare labeled conditions from the same session, and export a reproducible report. Native digital files are preferred; when only printouts exist, users may digitize with validated external tools[6–8] or use VectBox Sync’s optional PDF path for common manufacturer layouts.

This Technical article describes the technology, its development lineage, how it is used, its intended clinical and research role and limits, current problems, and planned developments.

## 2 History of development

We previously reported a VCG-based ventricular dyssynchrony index and a control-population cutoff for exploratory interpretation[2]. That analysis pipeline, implemented in MATLAB as well as in Python, already returned the index, companion VCG descriptors, and synchrony maps. In this article, we present VectBox Sync, a hosted browser application, to make the same method easier to use more widely without any installation. The application presented here is the first web version: it accepts native digital recordings and, optionally, manufacturer PDFs, and the analysis ends with a multilingual PDF report.

## 3 Description

### 3.1 Architecture and how it is used

VectBox Sync is a hosted web application that delivers an end-to-end VCG synchrony pipeline (Figure 1a): a browser single-page client at `/sync`, a FastAPI backend over HTTPS, and containerized deployment (Docker Compose with TLS termination). The backend orchestrates multi-format import, optional preprocessing, Kors-based VCG reconstruction, synchrony analysis, and PDF report generation. Uploaded files are processed in memory; they are not retained server-side in the default guest workflow. Session continuity may be kept locally in the browser under the user’s control.

**Figure 1.**
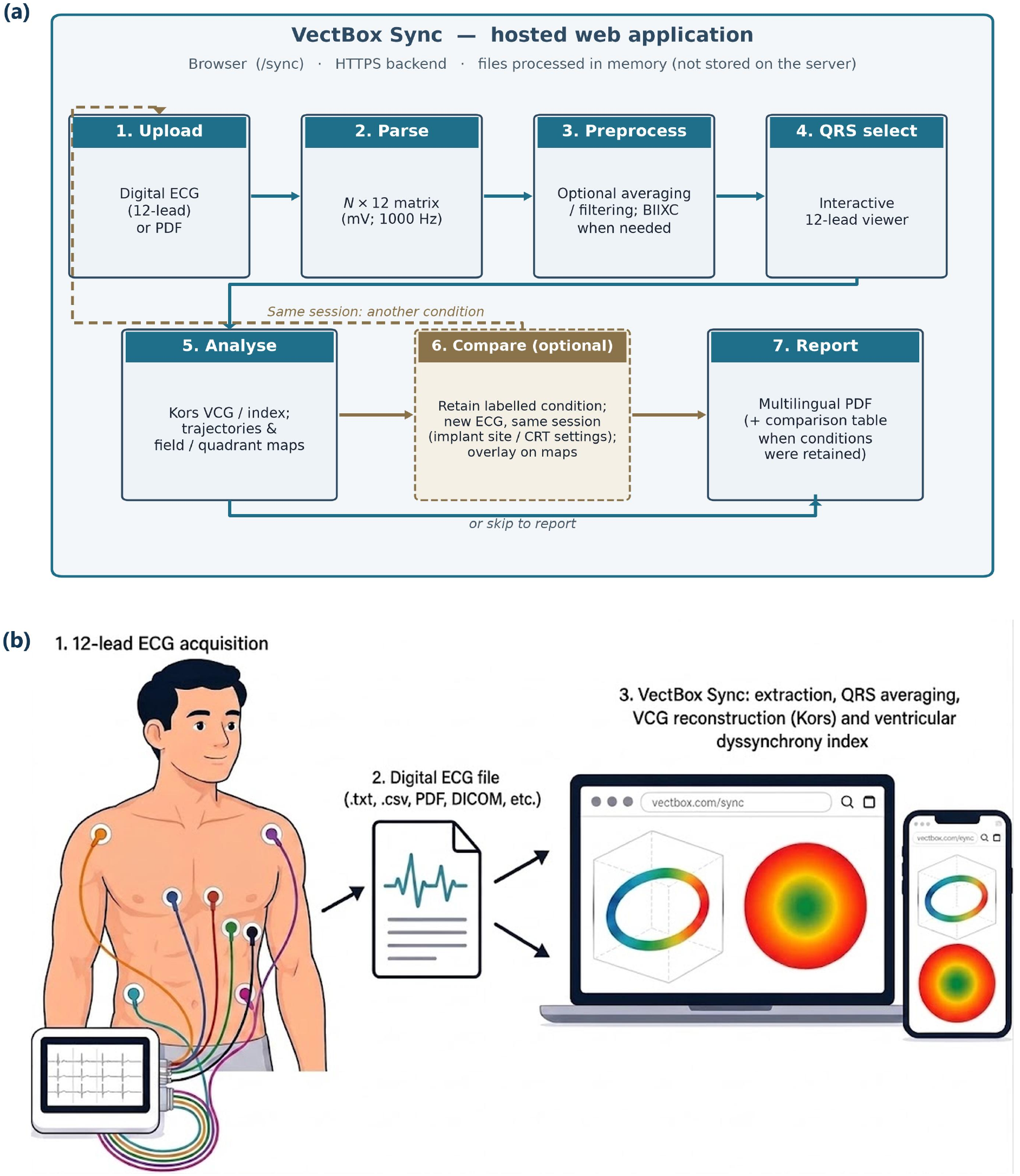
(a) End-to-end VectBox Sync pipeline in the hosted web application. Hosting is summarized in the top strip. Solid path: single-recording analysis. Dashed: optional within-session comparison of labeled conditions (for example CRT implant sites or device settings). (b) From 12-lead ECG acquisition to a digital file and browser analysis (extraction, optional QRS averaging, Kors VCG reconstruction, and the ventricular dyssynchrony index) on a laptop or phone at vectbox.com/sync.

Figure 1b shows the path from 12-lead ECG acquisition to a digital file and browser analysis on a laptop or phone. A typical session follows the steps in Figure 1a: (i) upload a digital ECG or optional manufacturer PDF; (ii) parse/extract to an *N* × 12 matrix (mV); (iii) optionally average beats and/or filter; (iv) select a QRS interval on an interactive 12-lead viewer; (v) compute the dyssynchrony index, companion VCG descriptors, trajectories, and synchrony-field maps; (vi) optionally retain the result as a labeled condition and repeat under another recording from the same session; (vii) download a multilingual PDF report. Native digital files are faster; a manufacturer PDF is the slowest input path and typically takes about 1–3 minutes from upload through report for a user already familiar with the interface. Atypical PDFs (raster scans or non-standard page geometry, for example several traces on one page) open an assisted grid-confirmation step that can prolong analysis time. The live application is at https://vectbox.com/sync.

### 3.2 Inputs

Preferred inputs already contain numerical samples (Table 1). All accepted files are normalized to a common internal matrix (*N* samples × 12 leads in mV; resampled to 1000 Hz when required). Matrices produced by external digitizers (`.txt, .csv`, or NumPy) are first-class inputs.

**Table 1.** Input formats accepted by VectBox Sync by the date of submission.

| Category | Extensions / encoding | Notes |
|---|---|---|
| Delimited matrix | .txt, .csv | *N* × 12 array |
| NumPy | .npy, .npz | NumPy arrays |
| Proprietary module export | .ecg | Native digital cart-module export |
| WFDB | .hea+.dat | PhysioNet-style |
| EDF / DICOM / SCP | .edf, .dcm, .scp | Clinical digital |
| Vendor / std. XML | .xml | HL7 aECG; GE MUSE; Philips Sierra |
| Manufacturer PDF | .pdf | Optional digitization |

Optional PDF loading routes among common layouts (Table 2) using vector-first extraction when possible. Embedded digital payloads (e.g. DICOM/XML attachments) are preferred over ink reconstruction. When a continuous full-width rhythm strip is present on a non-simultaneous grid, it is digitized with per-lead time offsets to enable BIIXC averaging (below). Photo-graph/screenshot digitization is out of scope; a quality-assurance (QA) overlay supports visual review of PDF extraction (Figure 2). When automatic layout detection is uncertain—raster PDFs or non-standard page geometry—an assisted extraction view lets the user confirm the layout and adjust the lead grid before analysis.

**Table 2.** Compatible PDF layouts.

| Layout | Simultaneous leads | Averaging |
|---|---|---|
| 12×1 | Yes | Optional |
| 6×2 | Yes | Optional |
| 3×4 | No | Required (BIIXC if a rhythm strip is present) |
| 6×2 sequential | No | Required (BIIXC if a rhythm strip is present) |

**Figure 2.**
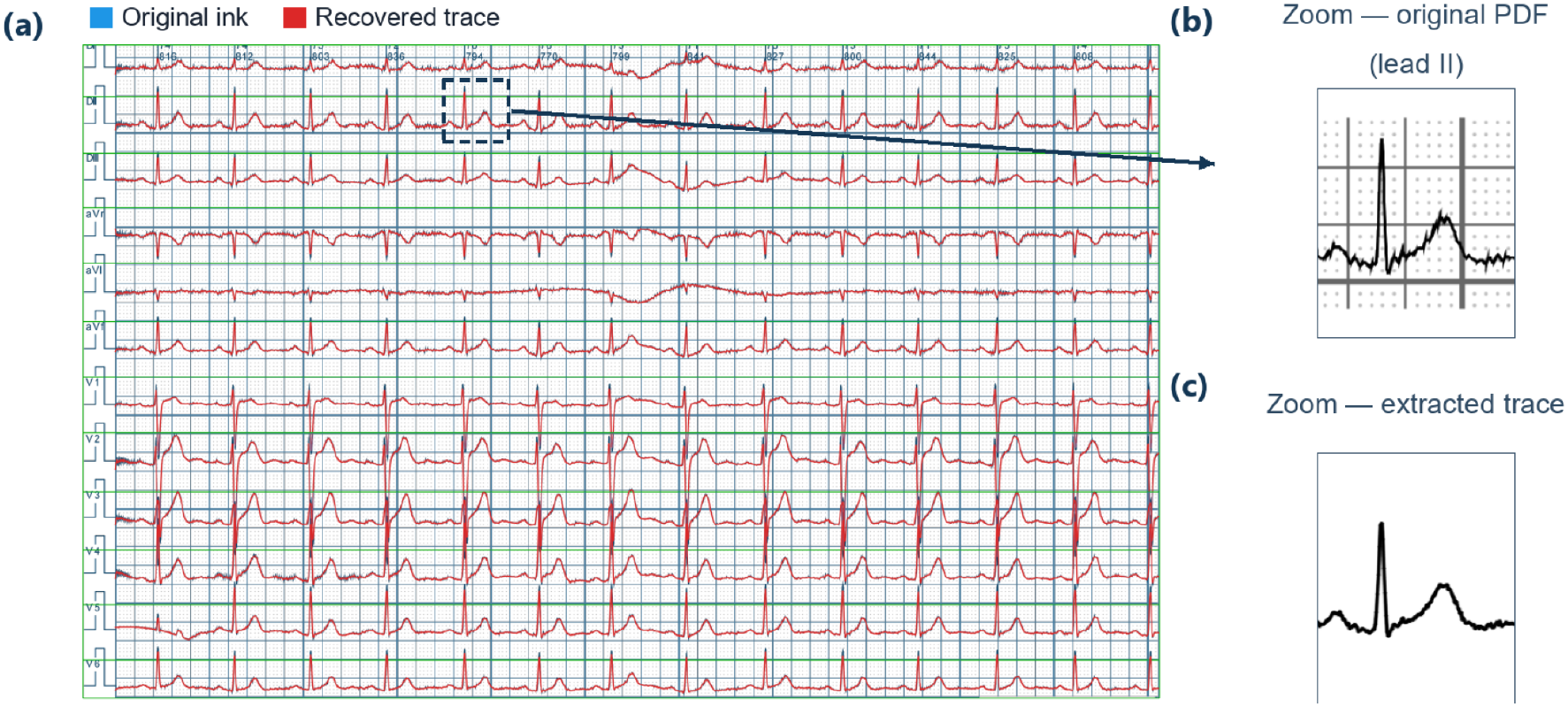
(a) QA overlay for PDF extraction from a vectorized 12×1 manufacturer PDF: blue marks the original ink and red the recovered trace. The rectangle marks the lead-II P–QRS–T window shown in (b) source PDF and (c) extracted polyline in the same pixel window without grid.

### 3.3 Preprocessing

On simultaneous recordings, optional beat averaging aligns each beat to the peak of the spatial vector magnitude (SVM), the instantaneous length of the three-dimensional cardiac vector. If that peak cannot be detected, alignment uses R peaks on lead II instead. An optional control on lead I allows users to include or exclude individual beats from the average. All detected beats are included by default, and any beat that would make the average unrepresentative can be left out. Pre-/post-R window lengths scale with median RR (approximately 0.38 RR before and 0.55 RR after R), capped to exclude neighbouring QRS complexes; the averaged beat is trimmed by default to the P–QRS–T triad plus short isoelectric margins.

On non-simultaneous printouts, averaging is required before analysis. If a usable rhythm strip is available, VectBox Sync applies BIIXC[9]; otherwise it falls back to per-lead averaging with quality gates (beat counts, active leads, template correlation). Optional filters include bandpass, baseline wander removal, mains notch, and respiration-band suppression.

### 3.4 Synchrony analysis and reporting

On a user-selected sample interval, VectBox Sync reconstructs XYZ leads with the Kors transform[10] and computes the published dyssynchrony index[2], returning the scalar index relative to the calibrated control cutoff, X–Y and X–Z projections (Figure 3a, top left and top right), vector magnitude and vector displacement speed (Figure 3a, bottom left and bottom right), two derived maps (Figure 3b), and a multilingual PDF report (English, Spanish, Portuguese, German, French, Italian, Chinese, and Russian). The same view and report also list companion QRS-loop descriptors: the voltage-time integral of spatial vector magnitude (VTI), analogous to QRS area[3, 4], and the speed-time integral of spatial vector displacement speed (STI), which are the coordinates from which the index is derived; frontal and horizontal VCG angles; and a planarity scalar (unity for a perfectly planar loop). These companion quantities are descriptive loop-geometry outputs, not additional validated CRT-response classifiers. Analysis is refused for non-simultaneous data that have not been averaged.

**Figure 3.**
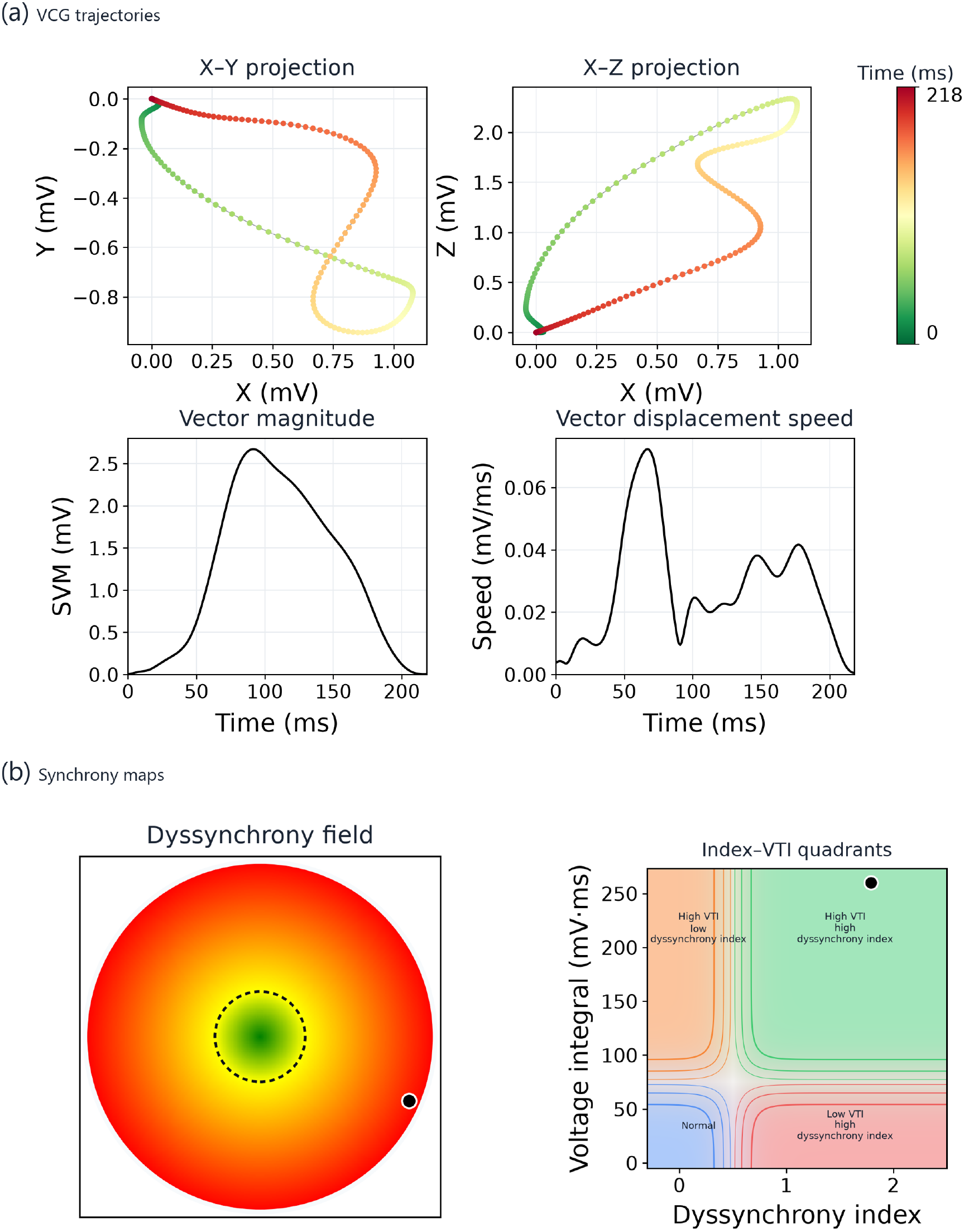
Representative Sync PDF-report panels from a de-identified clinical recording: (a) X– Y and X–Z projections, vector magnitude, and vector displacement speed over the selected QRS window. Color on the two projections encodes time within that window, from its onset (0 ms) to its end, and the magnitude and speed axes use that same millisecond scale (1000 Hz; one sample = 1 ms); (b) dyssynchrony field (bullseye) and index–VTI quadrants. In both maps the black marker is the patient’s location. Quadrant labels name high versus low VTI and high versus low index; they are exploratory descriptors, not a validated CRT-response classifier.

The dyssynchrony field is a polar (bullseye) view of the same index (Figure 3b, left panel): radius is the perpendicular distance from the patient’s voltage–speed (VTI–STI) coordinates to the control-population line, so the center corresponds to that relationship and larger radii to greater deviation. Color runs from green (below the cutoff) through yellow (at the cutoff) to red (above it); angular position encodes where the point lies along, and on which side of, the control line. In this framework the control line is the empirical expression of a conserved scale relationship (see Section 1). The index does not claim to measure delay between individual myocardial regions; it quantifies how far global ventricular electrical dynamics depart from the organized-normal VTI–STI relation[2]. A pure amplitude (scale) change should move a point along the line; a change in activation sequence should displace it off the line.

The quadrant map plots the index against VTI (Figure 3b, right panel). Index and VTI thresholds organize four exploratory regions, drawn as a continuous contour map rather than four hard rectangles, and labeled by high versus low VTI and high versus low index. The regions are descriptive, not a validated CRT-response classifier.

Within a session, successive analyses can be kept as labeled conditions and overlaid on the dyssynchrony field and quadrant maps, with a summary table in the PDF report. Intended exploratory uses include comparing left-ventricular lead sites or pacing vectors during CRT implant work-up, and comparing device settings after a CRT system has been implanted. The overlay displays electrical-synchrony differences across conditions; it does not prescribe lead placement or programming.

### 3.5 Technical validation

De-identified sample recordings were processed end to end to confirm that native digital files and optional manufacturer PDFs load across several formats and that, after QRS selection, the pipeline returns a finite dyssynchrony index and VCG outputs. Where averaging was used, averaging quality checks (beat counts, active leads, and template correlation) were also verified. As a secondary check of optional PDF loading, de-identified raw-data recordings were paired with manufacturer PDF exports of the same source in four common page layouts (12 × 1, 6 × 2, 3 × 4, and 6 × 2 sequential with a rhythm strip). After vector extraction, each PDF matrix was compared with its originating raw data (Pearson correlation and RMSE after subtracting each lead’s mean (demeaning), summarized as the median across the twelve leads). Agreement is summarized in Table 3.

**Table 3.** Waveform agreement between raw data and manufacturer PDFs (median across leads; PCC = Pearson correlation after subtracting each lead’s mean).

| Layout | *n* | Median PCC | PCC range | Median RMSE (mV) |
|---|---|---|---|---|
| 12×1 | 10 | 0.996 | 0.990–0.999 | 0.011 |
| 6×2 | 10 | 0.994 | 0.990–0.999 | 0.013 |
| 3×4 | 10 | 0.995 | 0.992–0.999 | 0.011 |
| 6×2 sequential | 10 | 0.997 | 0.995–0.998 | 0.012 |

## 4 Clinical role

Guideline-directed CRT selection still leaves a substantial non-responder fraction[1], and QRS morphology alone does not fully capture the spatial and temporal complexity of ventricular activation[2]. Two recordings can share similar QRS duration and/or QRS area yet differ in the space-time organization of the vector trajectory; our index is aimed at that organizational difference. Moreover, the same space-time organization of depolarization may not be equally effective in two hearts with different distributions of viable myocardium (e.g., ischemic scar). There is therefore a clinical research need for reproducible, quantitative electrical-synchrony measures that go beyond duration and pattern reading—for exploratory candidate characterization, teaching of spatial activation concepts, and hypothesis-generating comparisons with imaging or device data[2–5].

In practice, applying such VCG-derived indices usually requires local scripts or ad hoc analysis workflows. Open-source desktop packages can already take a digital 12-lead ECG through automated VCG construction and measurement[11]. VectBox Sync is a hosted, install-free delivery layer for the published dyssynchrony method[2]: centers can apply the same Kors-based workflow and reference cutoff without re-implementing reconstruction, QRS selection, maps, or reporting. It complements, rather than replaces, standard ECG interpretation, imaging-based mechanical dyssynchrony assessment, device interrogation, and other electrical-dyssynchrony markers studied in CRT populations, including QRS area[3, 4] and related VCG measures[5]. Relative to dedicated digitization systems[6–8], its contribution is not superior ink recovery but end-to-end synchrony analysis on digital inputs, with PDF import only as a convenience path.

Intended uses include standardized multicenter application of the index, VCG synchrony teaching, exploratory contrasts with QRS duration, morphology, or imaging markers, and within-session comparison of pacing conditions (candidate left-ventricular sites or post-implant optimization). Broader clinical adoption will require prospective outcome studies and external validation beyond this Technical article. VectBox Sync is a research and teaching aid; it does not replace clinical judgment.

## 5 Current problems

Several limitations currently affect the applicability of the software and will need to be addressed in future work. The scientific validity of the dyssynchrony index rests on the prior publication[2]; this article does not redefine the metric or report new clinical outcomes, and external validation of the web-computed index in independent cohorts remains outstanding. More generally, VCG amplitude- and area-based parameters can be sensitive to heart–torso geometry and electrode placement[12]. The Sync dyssynchrony index, however, measures deviation from the control voltage–speed (VTI–STI) relationship rather than absolute amplitude or area alone. Under the scale argument above, a change in electrically involved myocardium or in volume-conductor gain should move a point along that line rather than off it; we previously argued that the resulting deviation is therefore likely relatively independent of non-synchrony factors such as cardiac mass and the electrical properties of non-cardiac intrathoracic tissue (volume-conductor effects)[2]. Absolute voltage- or area-derived quantities shown alongside the index should still be read with geometry and electrode-placement caveats in mind. Photograph or screenshot digitization is not supported. Raster manufacturer PDFs are handled by assisted grid confirmation rather than fully automatic vector extraction; very low-resolution scans may still fail, in which case users may obtain numerical data with validated digitizing tools and upload the resulting matrix[6–8]. On non-simultaneous printouts without a usable rhythm strip, averaging falls back to a per-lead strategy that does not restore inter-lead timing; BIIXC is used when the strip is available[9]. Closed-source distribution limits third-party code audit, mitigated by interactive review, optional QA overlays for PDF inputs, and the algorithmic description herein. The present interface is interactive and case-wise, and the hosted service does not provide longitudinal cloud archives by default; continuity relies on local browser storage and user-held PDF/CSV exports.

## 6 Future developments

Planned developments include broader clinical validation of the web-computed index against imaging and therapy-response endpoints, expansion of suite tools beyond Sync, and continued hardening of multi-format import for real-world digital ECGs. Prospective clinical studies correlating the index with CRT response and hard clinical outcomes will be essential to establish its clinical utility and assess its potential relevance to clinical guidelines.

## 7 Conclusion

VectBox Sync provides a web-based, end-to-end implementation of our previously reported VCG-based ventricular synchrony analysis for 12-lead ECGs[2], prioritizing native digital ECG inputs while also supporting manufacturer-provided PDFs and externally digitized waveform matrices. The application is available at https://vectbox.com/sync for research and educational use. Technical validation included end-to-end testing with de-identified sample recordings and assessment of waveform agreement between paired raw-data and PDF recordings (Table 3). Prospective clinical validation will be required to establish its performance in clinical settings.

## Data Availability

All data produced in the present study are available upon reasonable request to the authors

## Declarations

### Ethics approval

Not applicable: this manuscript describes software and technical validation on de-identified test recordings; no prospective human subjects data were collected for this study.

### Data availability

De-identified validation figures will be made available upon reasonable request or as supplementary material. Uploaded user recordings are not retained on the server.

### Software availability

VectBox Sync is accessible at https://vectbox.com/sync (suite landing: https://vectbox.com). The implementation is proprietary; algorithmic details are speci-fied in this article.

### Author contributions

R. Goldman: writing—review and editing; software testing. D. Spag-nuolo: writing—review and editing; electrophysiology consultation. O. Chara: conceptualization of the dyssynchrony index; writing—review and editing. J. Fernández: conceptualization of the dyssynchrony index; design and implementation of the analysis pipeline; writing—original draft. All authors approved the final manuscript.

## Declaration of generative AI and AI-assisted technologies in the manuscript preparation process

During the preparation of this work the authors used Cursor in order to revise the wording and organization of the manuscript. After using this tool, the authors reviewed and edited the content as needed and take full responsibility for the content of the published article.

